# Comparative effectiveness of mRNA-1273 versus protein-based NVX-CoV2705 vaccination on COVID-19-related outcomes among US insured adults during 2024–2025: a retrospective matched cohort study

**DOI:** 10.64898/2026.04.02.26350067

**Authors:** Amanda Wilson, Ekkehard Beck, Heather Hensler, Nevena Vicic, Keya Joshi, Emily Patry, Linwei Li, Jane Wang, Chris Clarke

**Author notes:** Corresponding author: Amanda Wilson, Moderna, 325 Binney St., Cambridge MA 02142.

## Abstract

**Background:** COVID vaccination with periodically updated compositions remains important as SARS-CoV-2 continues to circulate, cause disease, and evolve. Available COVID-19 vaccines in the 2024-2025 season differed by platform, including mRNA-1273, an mRNA-based vaccine, and NVX-CoV2705, a recombinant protein-based vaccine and antigen composition (KP.2-targeted and JN.1-targeted, respectively). There is limited head-to-head real-world evidence comparing the effectiveness of these different approaches to prevention of severe outcomes with COVID-19. We compared mRNA-1273 with protein-based NVX-CoV2705 in insured US adults vaccinated during the 2024–2025 season.

**Methods:** We conducted a retrospective matched cohort study in a large US claims database. Adults aged 18 years or older who received mRNA-1273 or NVX-CoV2705 between Aug 31, 2024 and Feb 28, 2025 were eligible. Recipients were exactly matched 2:1 on key demographic and clinical factors and then weighted with stabilized inverse probability of treatment weights. Outcomes were medically-attended COVID-19 and hospitalization with COVID-19 from day 7 after vaccination through up to 180 days of follow-up. We calculated comparative vaccine effectiveness (cVE) as 100 × (1−hazard ratio).

**Results:** Of 858,138 eligible mRNA-1273 recipients and 34,667 eligible NVX-CoV2705 recipients, 69,140 and 34,570, respectively, entered the matched cohort. Median (Q1, Q3) follow-up was 180 (163, 180) days for mRNA-1273 and 180 (162,180) for NVX-CoV2705. Medically attended COVID-19 occurred in 706 (1.02%) mRNA-1273 recipients and 512 (1.48%) NVX-CoV2705 recipients; adjusted cVE (95% CI) was 31.7% (23.4%, 39.1%). Hospitalization with COVID-19 occurred in 61 (0.09%) and 49 (0.14%) recipients, respectively; adjusted cVE (95% CI) was 40.7% (13.5%, 59.4%). In the 47,754 mRNA-1273 recipients matched to 23,877 NVX-CoV2705 recipients aged ≥65, adjusted cVE (95% CI) was 25.7% (15.4%, 34.8%) against medically-attended COVID-19 and 41.7% (14.3%, 60.4%) against hospitalization with COVID-19.

**Conclusions:** In this insured US adult population, mRNA-1273 demonstrated greater effectiveness against medically attended COVID-19 and hospitalization with COVID-19 than the protein-based NVX-CoV2705. These findings highlight the potential public-health importance of considering vaccine platform and variant selection when planning for upcoming seasons.

## Introduction

Despite increasing endemicity and accumulated population immunity, COVID-19 continues to cause substantial illness and severe outcomes in the US, with a disproportionate burden among older adults and those with underlying medical conditions.^1^ As protection from previous infection or vaccination wanes over time and the virus continues to evolve with the potential to escape pre-existing immunity, ongoing COVID-19 vaccination remains necessary.^2,3^

Each year, the circulating strains are monitored in the US to inform FDA’s Vaccines and Related Biological Products Advisory Committee (VRBPAC) in their task of recommending strain updates and updated strain compositions for vaccination in the upcoming season. During the timeframe leading up to the 2024-25 strain selection meeting, JN.1 had been the dominant strain since January, 2024.^4^ However, at the time of the meeting in June 2024, the circulating strains had become more complex with no clear dominance between JN.1 descendants such as KP.2 and KP.3. This resulted in the FDA recommending a JN.1-lineage–containing vaccine for the 2024–25 season, with a preference for KP.2 where feasible.^5^ The FDA subsequently authorized seasonal vaccines, including KP.2-targeted mRNA vaccines from Moderna and Pfizer-BioNTech and JN.1-targeted protein-based vaccine from Novavax ^6,7^. This created a setting in which concurrently available vaccines differed not only by platform but also by antigen composition.^8,9^

Updated seasonal vaccine formulations have been repeatedly shown to induce robust neutralizing antibody responses in pre-clinical and clinical studies, including cross neutralization against related strains.^10^ In addition, absolute effectiveness studies have consistently shown that updated COVID-19 vaccines continue to provide clinically meaningful protection against medically attended illness and severe outcomes.^11,12^ However, real-world comparative evidence remains essential because vaccine choice in clinical practice often involves both composition and platform considerations, especially when clinicians, patients, and policy makers weigh the relative performance of available COVID-19 vaccines to best inform their seasonal vaccination plan.

Whereas substantial head-to-head real-world evidence exists comparing different COVID-19 mRNA vaccines,^13–17^ such evidence comparing mRNA and protein-subunit vaccine is limited. Specifically, an Australian study on primary and first booster doses showed greater vaccine effectiveness of mRNA vaccines compared to NVX-CoV2373 for prevention of SARS-CoV-2 infection.^18^ Korean studies evaluating primary series and boosters reported non-statistically significant differences in severe COVID infections between the protein-based NVX-CoV2373 and the COVID-19 mRNA BNT162b2 vaccine.^19,20^ To our knowledge, no large claims-based study has directly compared an updated mRNA vaccine with NVX-CoV2705 vaccine recipients during the 2024–25 season in the US. We therefore compared COVID-19-related outcomes following vaccination with mRNA-1273 versus protein-based NVX-CoV2705 among insured US adults during the 2024–25 season.

## Methods

### Study design and data source

We conducted a retrospective matched cohort study using deidentified medical and pharmacy claims from Optum’s de-identified Clinformatics® Data Mart, which is derived from a database of administrative health claims for members of large commercial and Medicare Advantage health plans. The analysis used data from Aug 31, 2023, to May 3, 2025. Because the study used secondary, de-identified data without direct patient contact, informed consent was not applicable and institutional review board oversight was not required.

### Participants and exposure

Adults aged 18 years or older who received one dose of KP.2-targeted mRNA-1273 (Spikevax) or protein-based JN.1-targeted NVX-CoV2705 (Nuvaxovid) between Aug 31, 2024, and Feb 28, 2025, were eligible. A subpopulation of patients aged 65 or older was also defined. Individuals were required to have continuous medical and pharmacy enrolment during the 365-day baseline period (allowing a 31-day gap) and at least one medical or pharmacy claim during baseline.

We excluded individuals with missing, unknown, or conflicting age, sex, region, or insurance type at index; adults aged 65 years or older covered by commercial insurance; receipt of more than one COVID-19 vaccine on the index date; a documented COVID-19 medical encounter or COVID-19 treatment within 90 days before or on the index date; another COVID-19 vaccine within 60 days before index; or a recorded death date on or before index.

### Matching and statistical analysis

mRNA-1273 recipients were exactly matched without replacement to NVX-CoV2705 recipients in a 2:1 ratio on age group, sex, region, insurance type, baseline status for US CDC defined high-risk conditions for severe COVID-19,^21^ prior-season COVID-19 vaccination history, and week of vaccination. Baseline covariates considered for weighting included prior COVID-19 diagnosis timing, healthcare utilization, influenza vaccination or diagnosis, Charlson comorbidity score, and individual comorbidities that contribute to high-risk status.

Within the matched cohort, we estimated stabilized inverse probability of treatment weights (IPTWs) using multivariable logistic regression. Covariate balance was assessed with absolute standardized differences. We evaluated medically attended COVID-19 from day 7 after vaccination (the eighth day since vaccination) through a maximum of 180 days of follow-up, defined by ICD-10-CM code U07.1 in any care setting. We also evaluated hospitalization with COVID-19, defined as an inpatient admission with U07.1 in any diagnosis position. The U07.1 diagnosis code has been shown to have high inpatient accuracy and reasonable predictive value across care settings.^22,23^

Individuals were censored at the earliest of the outcome of interest, receipt of an additional COVID-19 vaccine, loss of enrollment, death, 180 days of follow-up, or end of data availability. COVID-19 occurring during days 0–6 after vaccination was treated as an early infection censoring event. Cox proportional hazards models were used to estimate crude and IPTW-weighted hazard ratios (HRs) with 95% confidence intervals (CI); comparative vaccine effectiveness (cVE) was reported as 100 × (1−HR).

Data extraction and processing were conducted using an internal SQL-based platform, with analyses performed in R 4.5.0.

## Results

### Study population

Among people vaccinated during the intake period, 1,156,441 received mRNA-1273 and 45,384 received NVX-CoV2705. Exact 2:1 matching yielded an analytic cohort of 69,140 mRNA-1273 recipients and 34,570 NVX-CoV2705 recipients (Figure 1). Nearly all eligible NVX-CoV2705 recipients were retained; matching selected a comparator-similar subset of mRNA-1273 recipients.

**Figure 1.**
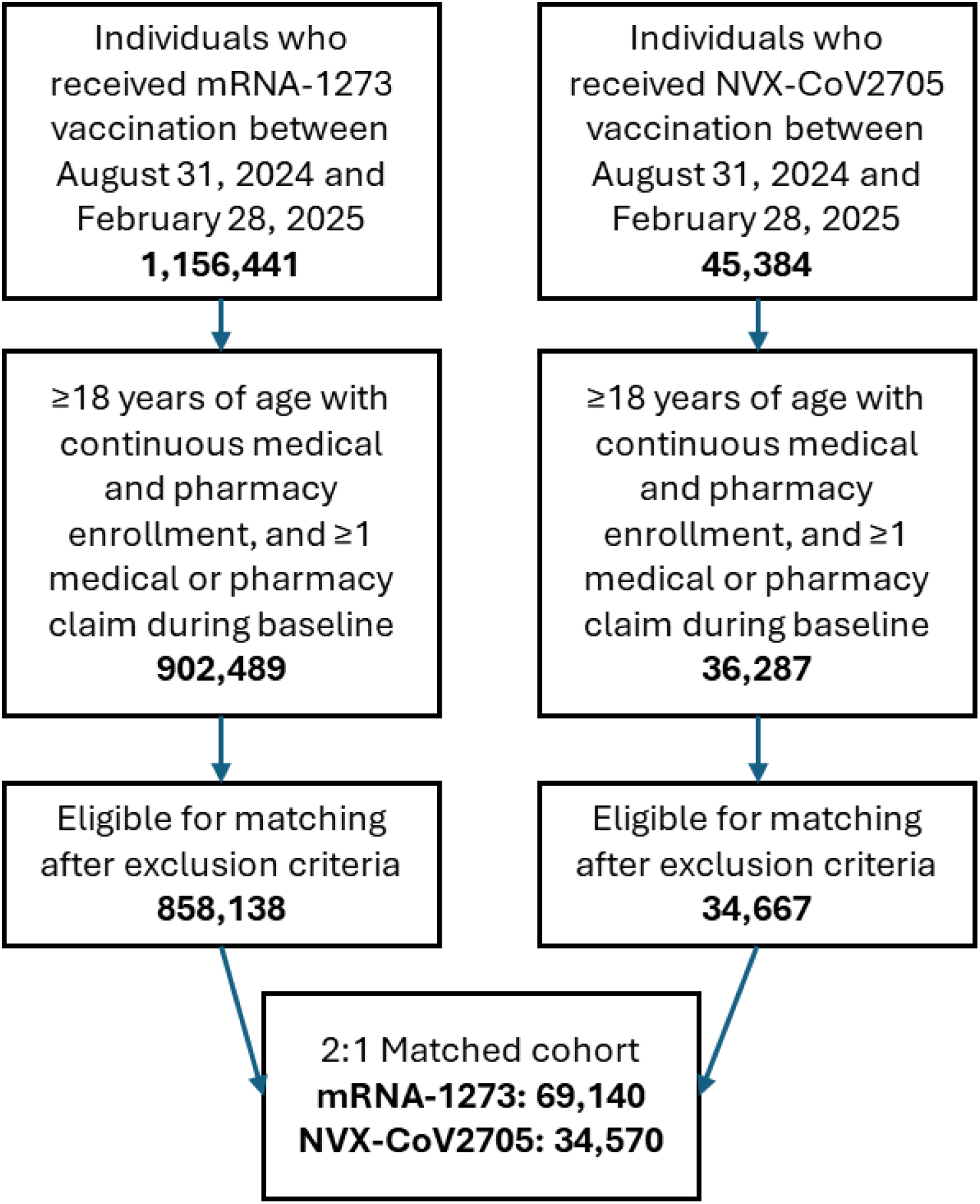
Study population.

The mean age of the matched cohort was 66 years; 69% of participants were aged 65 years or older; 57% were women; 71% had Medicare Advantage coverage; and 75% were classified as having at least one US CDC defined high-risk condition for severe COVID-19. Prior-season COVID-19 vaccination was recorded for 75% of both groups, and seasonal influenza vaccination for 81–82%. After weighting, all absolute standardized differences were 0.02 or lower, indicating excellent balance across measured covariates (Table 1).

**Table 1.**
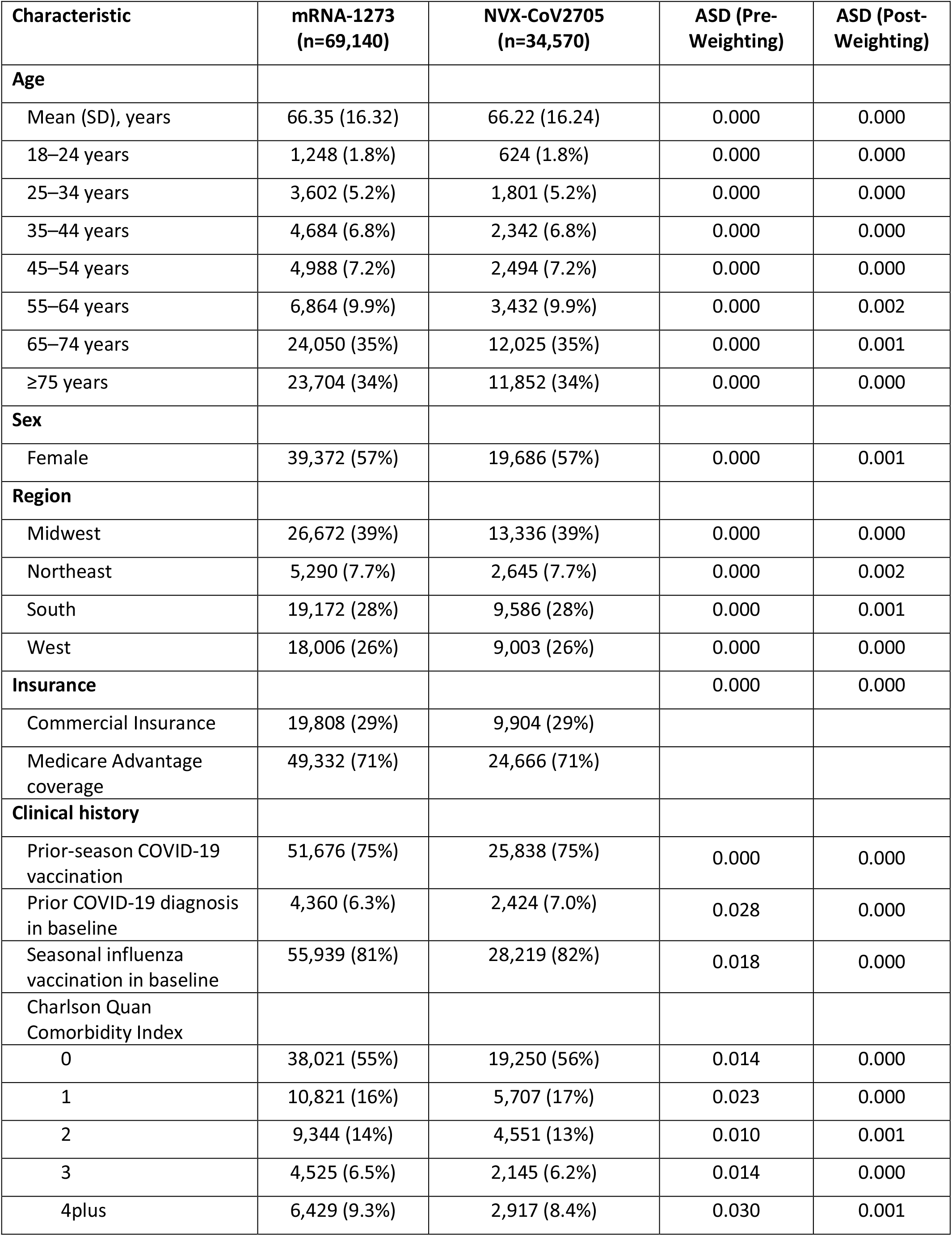
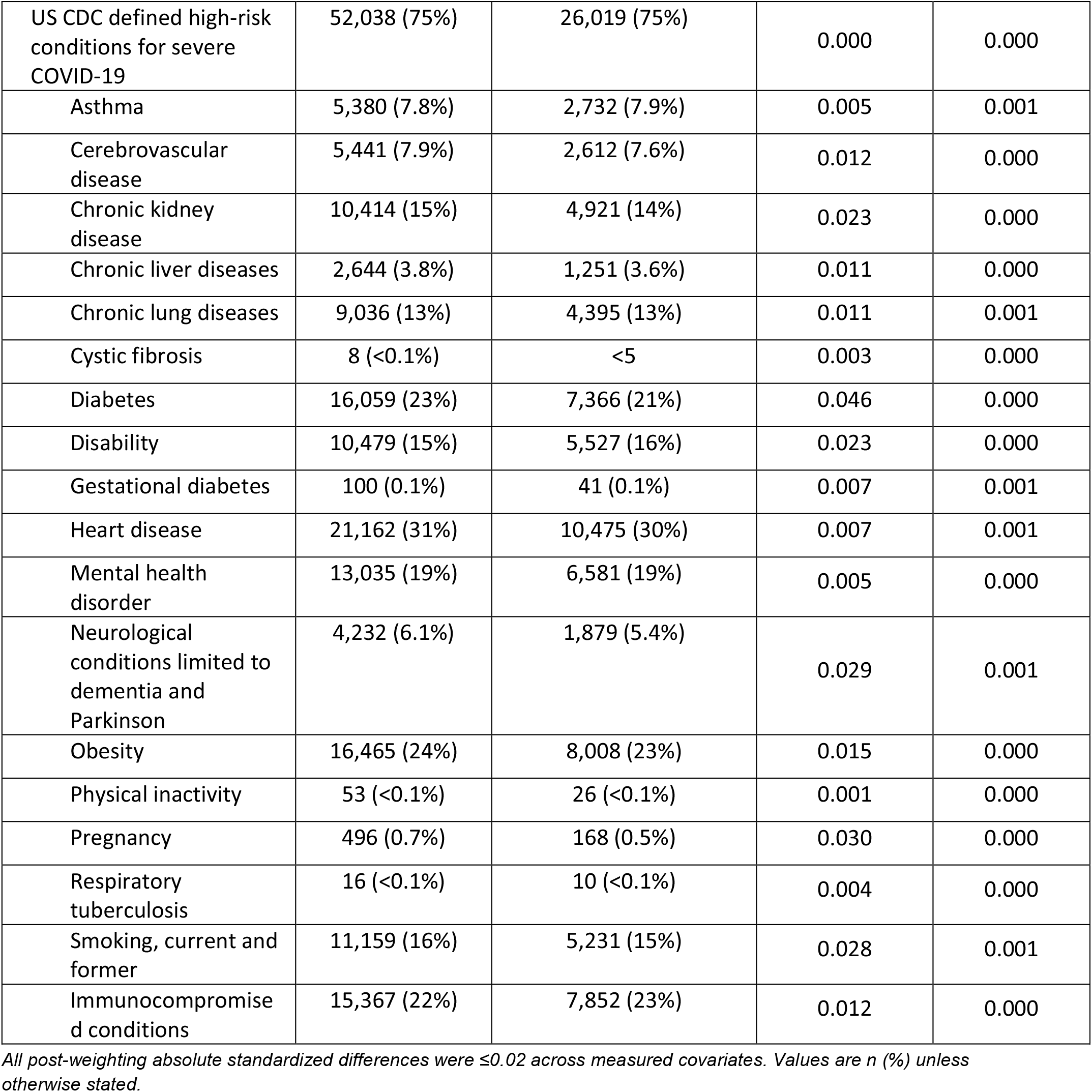
Selected baseline characteristics of the matched cohort after weighting.

| Characteristic | mRNA-1273<br>(n=69,140) | NVX-CoV2705<br>(n=34,570) | ASD (Pre-Weighting) | ASD (Post-Weighting) |
| --- | --- | --- | --- | --- |
| **Age** |  |  |  |  |
| Mean (SD), years | 66.35 (16.32) | 66.22 (16.24) | 0.000 | 0.000 |
| 18–24 years | 1,248 (1.8%) | 624 (1.8%) | 0.000 | 0.000 |
| 25–34 years | 3,602 (5.2%) | 1,801 (5.2%) | 0.000 | 0.000 |
| 35–44 years | 4,684 (6.8%) | 2,342 (6.8%) | 0.000 | 0.000 |
| 45–54 years | 4,988 (7.2%) | 2,494 (7.2%) | 0.000 | 0.000 |
| 55–64 years | 6,864 (9.9%) | 3,432 (9.9%) | 0.000 | 0.002 |
| 65–74 years | 24,050 (35%) | 12,025 (35%) | 0.000 | 0.001 |
| ≥75 years | 23,704 (34%) | 11,852 (34%) | 0.000 | 0.000 |
| **Sex** |  |  |  |  |
| Female | 39,372 (57%) | 19,686 (57%) | 0.000 | 0.001 |
| **Region** |  |  |  |  |
| Midwest | 26,672 (39%) | 13,336 (39%) | 0.000 | 0.000 |
| Northeast | 5,290 (7.7%) | 2,645 (7.7%) | 0.000 | 0.002 |
| South | 19,172 (28%) | 9,586 (28%) | 0.000 | 0.001 |
| West | 18,006 (26%) | 9,003 (26%) | 0.000 | 0.000 |
| **Insurance** |  |  | 0.000 | 0.000 |
| Commercial Insurance | 19,808 (29%) | 9,904 (29%) |  |  |
| Medicare Advantage<br>coverage | 49,332 (71%) | 24,666 (71%) |  |  |
| **Clinical history** |  |  |  |  |
| Prior-season COVID-19<br>vaccination | 51,676 (75%) | 25,838 (75%) | 0.000 | 0.000 |
| Prior COVID-19 diagnosis<br>in baseline | 4,360 (6.3%) | 2,424 (7.0%) | 0.028 | 0.000 |
| Seasonal influenza<br>vaccination in baseline | 55,939 (81%) | 28,219 (82%) | 0.018 | 0.000 |
| Charlson Quan<br>Comorbidity Index |  |  |  |  |
| 0 | 38,021 (55%) | 19,250 (56%) | 0.014 | 0.000 |
| 1 | 10,821 (16%) | 5,707 (17%) | 0.023 | 0.000 |
| 2 | 9,344 (14%) | 4,551 (13%) | 0.010 | 0.001 |
| 3 | 4,525 (6.5%) | 2,145 (6.2%) | 0.014 | 0.000 |
| 4plus | 6,429 (9.3%) | 2,917 (8.4%) | 0.030 | 0.001 |
| US CDC defined high-risk conditions for severe COVID-19 | 52,038 (75%) | 26,019 (75%) | 0.000 | 0.000 |
| Asthma | 5,380 (7.8%) | 2,732 (7.9%) | 0.005 | 0.001 |
| Cerebrovascular disease | 5,441 (7.9%) | 2,612 (7.6%) | 0.012 | 0.000 |
| Chronic kidney disease | 10,414 (15%) | 4,921 (14%) | 0.023 | 0.000 |
| Chronic liver diseases | 2,644 (3.8%) | 1,251 (3.6%) | 0.011 | 0.000 |
| Chronic lung diseases | 9,036 (13%) | 4,395 (13%) | 0.011 | 0.001 |
| Cystic fibrosis | 8 (<0.1%) | <5 | 0.003 | 0.000 |
| Diabetes | 16,059 (23%) | 7,366 (21%) | 0.046 | 0.000 |
| Disability | 10,479 (15%) | 5,527 (16%) | 0.023 | 0.000 |
| Gestational diabetes | 100 (0.1%) | 41 (0.1%) | 0.007 | 0.001 |
| Heart disease | 21,162 (31%) | 10,475 (30%) | 0.007 | 0.001 |
| Mental health disorder | 13,035 (19%) | 6,581 (19%) | 0.005 | 0.000 |
| Neurological conditions limited to dementia and Parkinson | 4,232 (6.1%) | 1,879 (5.4%) | 0.029 | 0.001 |
| Obesity | 16,465 (24%) | 8,008 (23%) | 0.015 | 0.000 |
| Physical inactivity | 53 (<0.1%) | 26 (<0.1%) | 0.001 | 0.000 |
| Pregnancy | 496 (0.7%) | 168 (0.5%) | 0.030 | 0.000 |
| Respiratory tuberculosis | 16 (<0.1%) | 10 (<0.1%) | 0.004 | 0.000 |
| Smoking, current and former | 11,159 (16%) | 5,231 (15%) | 0.028 | 0.001 |
| Immunocompromised conditions | 15,367 (22%) | 7,852 (23%) | 0.012 | 0.000 |

*All post-weighting absolute standardized differences were $\leq 0.02$ across measured covariates. Values are n (%) unless otherwise stated.*

### COVID-19-related outcomes

During follow-up, medically attended COVID-19 occurred in 706 of 69,140 mRNA-1273 recipients and 512 of 34,570 NVX-CoV2705 recipients. Median follow-up (Q1, Q3) was 180 (163, 180) days for mRNA-1273 and 180 (162,180) days for NVX-CoV2705. Cumulative incidence was 1.02% (0.95%,1.10%) for mRNA-1273 and 1.48% (1.35%, 1.61%) for NVX-CoV2705. The adjusted cVE (95% CI) was 31.7% (23.4%, 39.1%). In the population aged ≥65, there were 565 events in the 47,754 patients in the mRNA-1273 group and 379 events in the 23,877 patients NVX-CoV2705 group; adjusted cVE (95% CI) was 25.7% (15.4%, 34.8%) against medically-attended COVID-19.

Among all adults, hospitalization with COVID-19 was less frequent, with 61 events among mRNA-1273 recipients and 49 events among NVX-CoV2705 recipients. Cumulative incidence was 0.09% (0.07%, 0.11%) for mRNA-1273 and 0.14% (0.10%, 0.18%) for NVX-CoV2705. The adjusted cVE was 40.7% (13.5%, 59.4%), corresponding to an adjusted HR of 0.59 (0.41, 0.87). In the population aged ≥65, there were 56 events in the mRNA-1273 group and 48 events in the NVX-CoV2705 group; adjusted cVE (95% CI) was 41.7% (14.3%, 60.4%) against hospitalization with COVID-19. (Table 2).

**Table 2.**
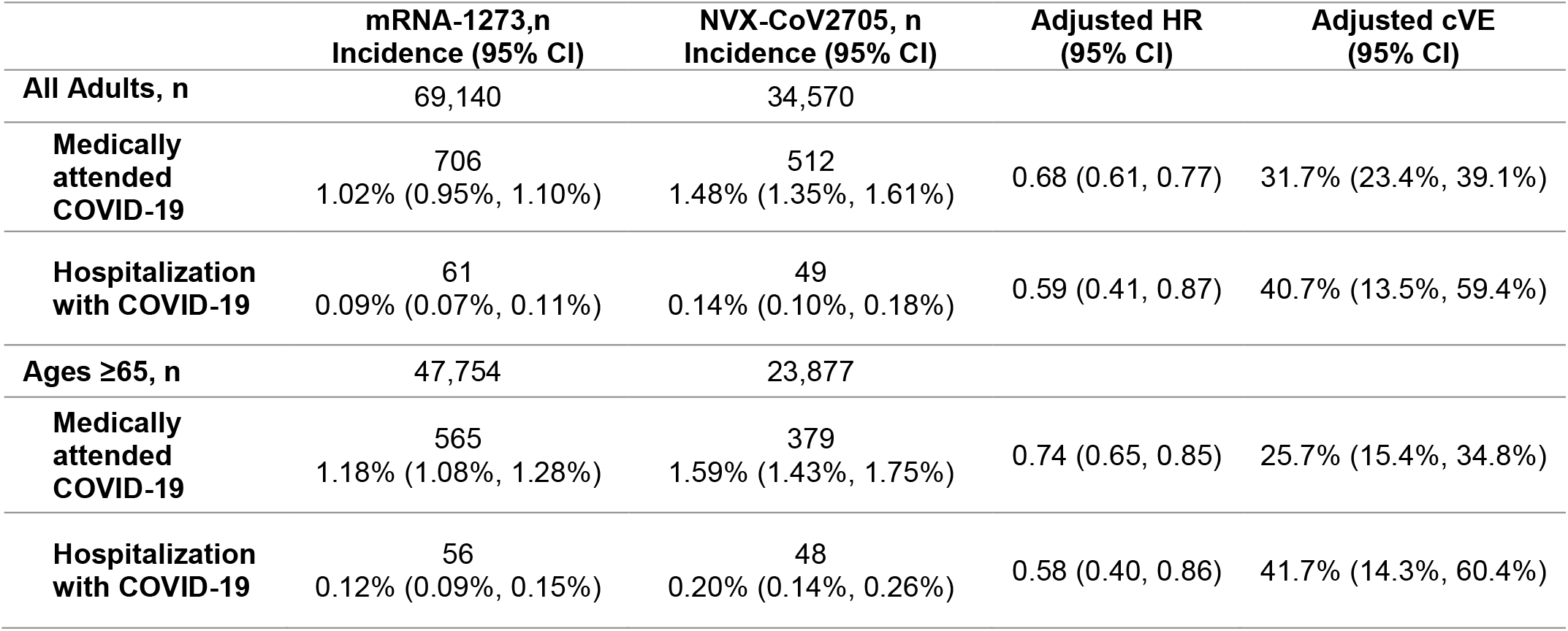
Comparative vaccine effectiveness of COVID-19-related outcomes.

The cumulative incidence curves separated early after vaccination and remained distinct throughout follow-up, with no evidence of attenuation in effect over time (Figure 2 and Figure 3). Most censoring reflected administrative end of follow-up or end of study (about 85% in each group) or loss of enrolment (about 12% in each group); receipt of another COVID-19 vaccine during follow-up occurred in less than 1.5% of participants.

**Figure 2.**
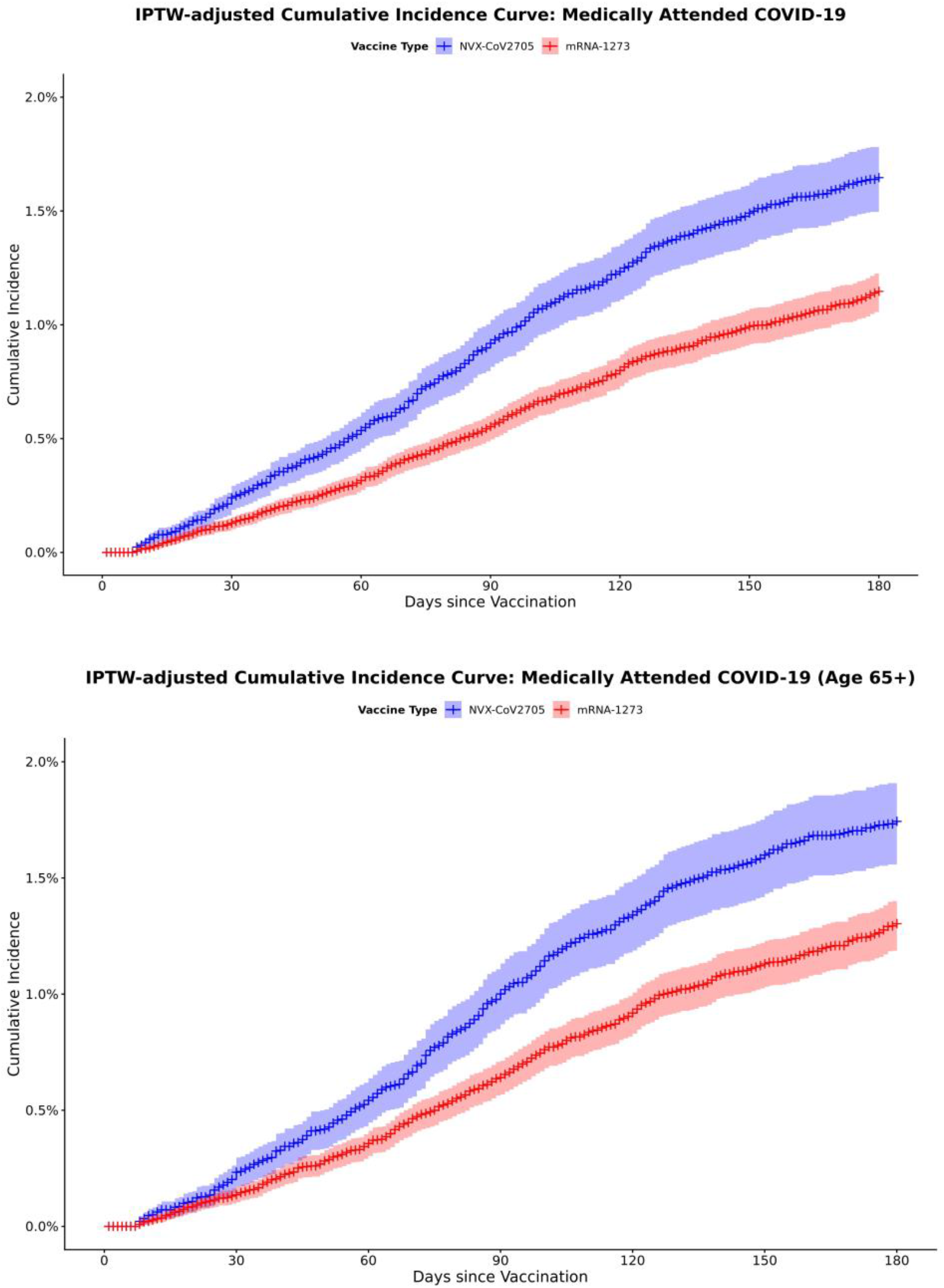
Cumulative Incidence Curves: Medically Attended COVID-19.

**Figure 3.**
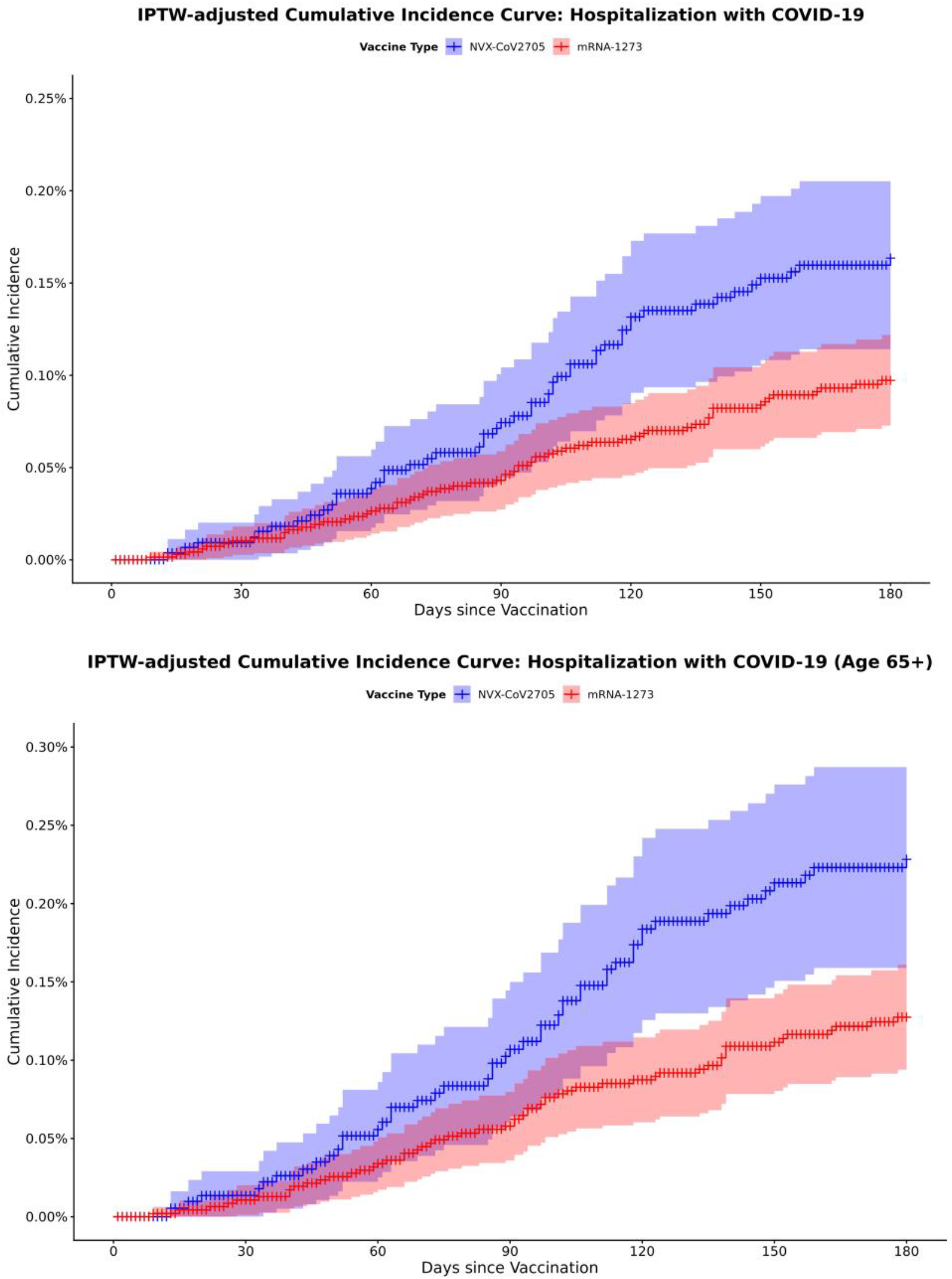
Cumulative Incidence Curves: Hospitalization with COVID-19.

## Discussion

In this large, observational, real-world study of insured US adults vaccinated during the 2024-2025 season, mRNA-1273 was associated with a lower risk of medically-attended COVID-19 and hospitalization with COVID-19 than the protein-based NVX-CoV2705, with a cVE of 31.7% (95%CI 23.4, 39.1) and 40.7% (95%CI 13.5, 59.4), respectively. This difference was reported over a median of approximately six months of follow-up, equivalent to the duration of a complete respiratory season. The early and sustained separation of cumulative incidence curves suggest persistent protection over the follow-up period. Considering the difference in cumulative incidence of medically-attended COVID-19, including hospitalization with COVID-19, vaccinating with mRNA-1273 is likely to translate to a substantial additional public health benefit.

Our study adds to the extensive knowledge base evaluating real world effectiveness of COVID vaccines, which has been of substantial value in confirming outcomes of randomized trials and informing public health policy during the transition to endemic phase. This study extends the limited evidence comparing mRNA and protein-based COVID-19 vaccines. Comparative vaccine effectiveness provides a complementary approach that helps decision makers interpret product-level performance when several updated vaccines are recommended concurrently.^12,24,25^

Several factors may underlie the observed differences between vaccines in this study. In particular, the mRNA and recombinant protein vaccines assessed differ in both mechanism of action and antigen target, with either or both of these factors potentially contributing to the observed differences.^26^

First, differences in mechanism of action may help explain the comparative VE observed in this study. Although this study was not designed to address immunological questions, these findings offer some insights into the clinical relevance of previous immunological comparisons between protein and mRNA COVID-19 vaccines. The higher VE observed for mRNA-1273 in this study is consistent with observations in head-to-head studies that show nAb and CD8 T cell responses are more potent for mRNA vaccine than protein vaccine.^27–29^ These findings also provide context for the potential role of IgG4 in vaccine-induced immunity to COVID-19. IgG4 antibody levels have been measured following repeated vaccination with mRNA vaccines,^30^ although their clinical significance remains uncertain. The higher VE observed for mRNA-1273 in this study does not support the hypothesis that increased IgG4 antibody levels are associated with reduced protection and is consistent with previous findings that antibody produced in response to mRNA-1273 has similar Spike protein neutralizing activity and retains Fc-mediated effector functions.^30^

Second, differences in antigen target may also have contributed to the comparative VE observed in this study. VE is known to be influenced by the extent of match between the vaccine antigen and circulating strains.^31,32^ In this context, mRNA-1273 targeted KP.2, whereas NVX-CoV2705 targeted JN.1. Meanwhile, SARS-CoV-2 epidemiology evolved rapidly over the season: KP.3 lineage viruses predominated in the fall, XEC became established during winter, and LP.8.1 emerged in early spring.^33^ Although JN.1 and KP.2 are closely related, both preclinical and clinical data indicate that KP.2-adapted vaccines elicit higher neutralizing antibody responses against a broader range of emerging JN.1-lineage variants compared with JN.1-based formulations.^34^ This suggests the potential for incremental protection with KP.2-targeted vaccines in a dynamically evolving variant landscape.

Taken together, these findings may reflect, in part, how the inherent platform flexibility and shorter manufacturing timelines of mRNA technology could provide public health benefits. In addition to the clinical insights of better immune response with mRNA-1273 as compared with a protein-based vaccine, the mRNA platform allows for later strain selection, enabling closer alignment between vaccine antigens and circulating strains at the time of vaccination campaigns, and potentially contributing to improved VE. However, as our estimates capture overall comparative performance rather than isolate a platform effect, further research is needed to distinguish the factors contributing to the observed comparative vaccine effectiveness.

Strengths of this study include the same-season head-to-head design, use of a large, well-validated US claims database, exact matching on calendar time and key confounders, and additional weighting across a broad set of measured covariates. The medically attended COVID-19 endpoint captures events leading to healthcare contact, and the endpoint of hospitalization with COVID-19 is less-subjective to healthcare seeking behavior and reflects severe COVID-19 outcomes, making it a policy-relevant endpoint.

This study also has limitations. As in all observational studies, residual confounding from unmeasured factors such as healthcare-seeking behavior, frailty, occupational exposure, or home testing practices might remain. Outcomes were claims-based and did not require laboratory confirmation within the data source; although U07.1 performs well in inpatient data and reasonably in mixed settings, some misclassification may occur, particularly for outpatient encounters.^22,23^ The hospitalization endpoint might include incidental SARS-CoV-2 infection recorded during an admission for another reason, however the approach is consistent with similar studies.^13,35,36^ Overall, the comparator uptake was low in the dataset, so only a subset of eligible mRNA-1273 recipients entered the exact-matched cohort, which improves exchangeability but narrows generalizability. Nevertheless, the demographics of the mRNA-1273-vaccinated cohort were not remarkably different to those reported in studies of absolute vaccine effectiveness, indicating the population may be generalizable to the larger vaccinated population.^35^ In addition, the database does not represent uninsured populations or traditional fee-for-service Medicare users, which affects generalizability.

## Conclusion

In conclusion, among insured US adults vaccinated during the 2024–2025 season, mRNA-1273 demonstrated greater effectiveness against medically attended COVID-19 and hospitalization with COVID-19 than the protein-based NVX-CoV2705 over up to 180 days of follow-up. These results add head-to-head US evidence on the comparative performance of updated mRNA versus protein-based COVID-19 vaccines in routine care and support prioritizing updated vaccine compositions, even in scenarios where prior-season vaccines remain available while variant selection for the upcoming season is still being determined. Replication in other datasets and in specific high-risk subgroups will be important. These findings support continued comparative evaluation of updated COVID-19 vaccines as product-level differences remain relevant for clinical and policy decision making.

## Data Availability

The study used deidentified administrative claims data licensed from Optum®. Individual-level data are not publicly available. Deidentified aggregate data may be shared, subject to sponsor review, data-provider permissions, and an appropriate data-use agreement.

## Funding

Moderna, Inc

## Declaration of interests

Authors are employees and stockholders of Moderna, Inc.

## Acknowledgments

The authors acknowledge that artificial intelligence was used in the drafting of the manuscript. All text and references were checked and revised by authors.

## Supplemental Materials

**Supplemental Table 1.** Censoring Reasons.

|  | **mRNA-1273** | **NVX-CoV2705** |
| --- | --- | --- |
| **Medically-attended COVID-19** |  |  |
| Another COVID-19 vaccination reported | 664 (0.96%) | 462 (1.34%) |
| Death | 600 (0.87%) | 277 (0.80%) |
| Early infection (<day 7) | 25 (0.04%) | 23 (0.07%) |
| End of study | 58743 (84.96%) | 29206 (84.48%) |
| Event within follow-up period | 706 (1.02%) | 512 (1.48%) |
| Loss of enrollment | 8402 (12.15%) | 4090 (11.83%) |
| **Hospitalization with COVID-19** |  |  |
| Another COVID-19 vaccination reported | 677 (0.98%) | 471 (1.36%) |
| Death | 603 (0.87%) | 279 (0.81%) |
| Early infection (<day 7) | 25 (0.04%) | 23 (0.07%) |
| End of study | 59334 (85.82%) | 29625 (85.70%) |
| Event within follow-up period | 61 (0.09%) | 49 (0.14%) |
| Loss of enrollment | 8440 (12.21%) | 4123 (11.93%) |

## Notes

### Author Declarations

This is a secondary analysis of de-identified data without access to personal identifying information and without direct enrollment of patients (e.g., in contrast to interventional studies or other primary data collection studies requiring patient participation). Thus, informed consent is not applicable. All patient-level and provider-level data within the database contain synthetic identifiers to protect the privacy of patients and data contributors. All study reports will contain aggregated data only that cannot be used to identify patients or physicians. Analysis data sets are Health Insurance Portability and Accountability Act (HIPAA) compliant. Per 45 CFR 46.104 d4, Institutional Review Board (IRB) approval is not required for secondary research, which does not use any identifiable information and if the study team does not have the code key with which the information can be made identifiable. This study is therefore not considered human subjects research and exempt from requiring IRB oversight.

